# A reappraisal of polymyxin B dosing based on population pharmacokinetic model in patient with renal insufficiency

**DOI:** 10.1101/2020.01.24.20018481

**Authors:** Xuben Yu, Chunhong Zhang, Jingye Pan, Ying Dai, Ziye Zhou, Fan Yang, Ruolan Sun, Yexuan Wang, Yancheng Cao, Changcheng Sheng, Zheng Jiao, Guanyang Lin

**Affiliations:** Department of Pharmacy, the First Affiliated Hospital of Wenzhou Medical University, Wenzhou, China; Department of Pharmacy, Shanghai Chest Hospital, Shanghai Jiao Tong University, Shanghai, China; Department of Pharmacy, Wenzhou Medical University, Wenzhou, China; Department of Pharmacy, Huashan Hospital, Fudan University, Shanghai, China

**Keywords:** polymyxin B, pharmacokinetics, dose individualization

## Abstract

**Background:** Current FDA-approved label recommends polymyxin B dosing should be adjusted according to renal function, despite several studies proved poor correlation between polymyxin B PK and creatinine clearance. The study aims to assess the impact of renal function on polymyxin B metabolism and identify an alternate dosing strategy by population analysis.

**Methods:** Blood samples from adult patients were collected at steady state during routine therapeutic drug monitoring. Nonlinear mixed effects modeling was employed to build a population PK model of polymyxin B. Monte Carlo simulations were performed to design polymyxin B dosing regimens across various renal function.

**Results:** Pharmacokinetic analyses included 112 polymyxin B concentrations at steady state from 32 adult patients aged 37-93 received intravenous polymyxin B (100-200 mg/d). The creatinine clearance in patients was 5.91-244 mL/min. In the final population PK model, CrCL was the significant covariate on CL (typical value, 1.59 L/hr; between-subject variability, 13%). Mean (SD) individual empirical Bayesian estimates of CL was 1.75 (0.43) L/hr. A new dosing strategy combining the PK/PD targets and Monte Carlo simulation indicated that polymyxin B dose reductions improved the probability of achieving optimal exposures in simulated patients with renal insufficiency. For severe infections caused by organisms with MIC of ≥ 2 mg/L, though a high daily dose (e.g. 200mg/day) would possible for bacterial eradication, the risk of nephrotoxicity is significantly increased.

**Conclusion:** A population PK model was established to develop individualized polymyxin B dosage regimens that the dose of polymyxin B should be adjusted according to CrCL.

## Introduction

Polymyxins (polymyxin B and colistin) become available clinically in the 1950s to treat Gram-negative bacterial infections. However, they were fell out of favor in 1970s due to the narrow therapeutic window and their potential to cause nephrotoxicity and neurotoxicity [1,2]. Over the last decade, polymyxins are again being used due to the emergence of multidrug-resistant (MDR) Gram-negative bacteria, combined with a paucity of new antibiotics [3-6]. Polymyxins, include polymyxin B and polymyxin E (colistin), are available for clinical use. Colistin is administered parenterally in the form of an inactive prodrug (colistin methanesulfonate, CMS) [7]. In contrast, polymyxin B is administered parenterally as its active form, which leads to less complex pharmacokinetics. Therefore, polymyxin B has been increasingly used due to its active form and potentially less nephrotoxic than colistin [8,9].

The clinical PK/PD of polymyxin B are scarce. Only a very small number of studies have examined the pharmacokinetics of polymyxin B following parenteral administration in human. Polymyxin B is not absorbed from the normal alimentary tract. The drug is excreted slowly by the kidneys. Its tissue diffusion is poor and the drug does not pass the blood brain barrier into cerebrospinal fluid. It should be noted that the PK data in the product information of polymyxin B for injection were obtained from studies conducted more than 40 years ago. The recent studies focused on PK analysis of polymyxin B pointed out that it was not significantly eliminated by the kidney and polymyxin B total body clearance was poorly correlated with creatinine clearance [10-11]. Therefore, the dosage of polymyxin B was not necessarily to be adjusted in patients with renal insufficiency according to the PK rationale [12]. However, current FDA-approved prescribing information recommends dose reducing “downward for individuals with renal insufficiency” [13], but the specific dosing strategy for patients with renal insufficiency is not provided by the package inserts. Although the recent reports on the PK of polymyxin B refute this recommendation, these studies were limited with a small number of samples. Therefore, larger PK studies in patients with renal insufficiency are needed to validate the recommendations provided by the package insert for polymyxin B and optimize its clinical use in patients with renal insufficiency. In the current study, we aim to identify the impact of patient renal function on polymyxin B metabolism and determine pragmatic dose modifications for patients with renal insufficiency using real-world data from routine therapeutic drug monitoring (TDM).

## Patients and Methods

### Patients and Ethics

The study was approved by the Ethical Committees of the First Affiliated Hospital of Wenzhou Medical University ([2019]034), China. From June 2018 to May 2019, adult patients (≥18 years) treated with intravenous polymyxin B for more than 48 hours with at least one steady-state polymyxin B serum concentration measured were included. Patients who died within 24 hours after the use of polymyxin B were excluded from this study. The consent were free passed by Ethics Committee in Clinical Research (ECCR)of the First Affiliated Hospital of Wenzhou Medical University.

### Data sources

Polymyxin B PK data were obtained retrospectively from a database maintained at the Department of Pharmacy reflecting routine clinical TDM in 32 patients (aged ≥ 18 years) treated at the First Affiliated Hospital of Wenzhou Medical University. Per protocol, blood samples were collected after ≥ 48 hours of treatment with polymyxin B (polymyxin B sulfate; for Injection). Polymyxin B C_max_ is obtained 30 minutes after the end of intravenous infusion, while C_min_ is obtained 30 minutes prior to the next dose. Dates and exact time of polymyxin B treatment and TDM were able to be indexed. The quantification of plasma concentration was performed using a validated high performance liquid chromatography-tandem mass spectrometry (LC-MS/MS) assay [14]. Intra- and inter-day assay coefficients of variation were <10% and the lower limit of quantification was 0.1 mg/L. Demographic data, intensive care unit admission, serum creatinine measurements, and other laboratory values were available in the database. Estimated creatinine clearance was calculated using the Cockcroft-Gault equation. Data organization and visualization was performed using R (version 3.6.0) and R Studio (version 1.2.1335).

### Pharmacokinetic Analysis

Population PK analysis was performed using non-linear mixed-effects modeling in NONMEM (version 7.4, Icon Development Solutions, Ellicott City, MD, USA) implemented using Pirana (version 2.9.7). R (version 3.6.0) and Xpose (version 4.3.2) software packages were used to generate diagnostic plots. The first-ordered conditional estimation method with inter- and intra-subject variability was used throughout the model development process.

One- and two-compartment models were explored for the polymyxin B serum concentration-time profiles with linear or nonlinear elimination. Fixed effects were parameterized in terms of CL and V. Between-subject variability (BSV) was modeled using exponential function. Residual variability was assessed using additive, proportional, and combined (additive plus proportional) error models. The base model was selected based on the Akaike Information Criteria (AIC) and visual inspection of diagnostic plots.

Exploratory analysis was performed before covariate modeling to examine the distribution of covariates in the population as well as correlation between covariates of interest. Primary covariates included demographics (age, sex), body size (total body weight [TBW]), creatinine clearance (CrCL), serum albumin concentration. Relationships between individual empirical Bayesian estimates of PK parameters and patient covariates were examined visually. Covariates were included using a forward selection process with a threshold decrease in OFV of 3.84 (p < 0.05, 1 degree of freedom [df]) until no further decrease in OFV was observed. In backward elimination, the covariate was retained in the final model with a threshold increase in OFV of 6.63 (p<0.01, 1 df), otherwise, it was eliminated from the model. The additional criterion for retaining the covariate in the final model was a decrease in the unexplained BSV and increase in PK parameter estimate precision [15].

For model evaluation, diagnostic plots were created to evaluate the final model including observed versus individual-fitted and observed versus population-fitted polymyxin B concentrations, conditional weighted residuals versus population predicted concentration and time since last dose. To evaluate the predictive performance, the statistics of the observed and simulated time-concentration profiles were compared using prediction- and variability-corrected visual predictive check (pvcVPC) [16] and normalized prediction distribution error (NPDE) test [17]. The dataset was simulated 1000 times using the $SIMULATION block in NONMEM® for pvcVPC and NPDE.

### Monte Carlo Simulations of Dosage Regimens

Monte Carlo simulations were performed using the final model to identify pragmatic potential polymyxin B dose adjustments. Total daily doses of 60, 80 100, 150, 200 mg with 12 hours interval were evaluated. Target values of AUC_ss, 24hr_ between 50-100 mg·hour/L was recommend by the current guidelines for the optimal use of polymyxins [12]. Probabilities of target attainment were calculated for each dosing simulated for 5000 virtual subjects with creatinine clearance ranging from 10 to 120 mL/min.

## Results

### Baseline characteristics of patients

Demographic data of the 32 adult patients are presented in Table 1. A large range of renal functions was observed. The CrCL were ranged from 9.12 to 146.7 mL/min in 29 patients, and 3 patients was with extremely lower serum creatinine below 35 μmol/L. The dose of polymyxin B was 100-200 mg/day. Most of the patients were received the drug every 12 hours.

**Table 1.** Patient Characteristics.

| Characteristic | Value<sup>a</sup> |
| --- | --- |
| Age (years) | 63.63 (12.92) |
| Sex |  |
| Male | 26 (82.25%) |
| Female | 6 (18.75%) |
| Height (cm) | 167.97 (6.69) |
| Total body weight (kg) | 61.73 (11.77) |
| CrCL (mL/min) | 86.58 (53.00) |
| Serum albumin (g/L) | 30.15 (5.76) |
| Serum creatinine (μmol/L) | 116.69 (127.60) |

Abbreviations: CrCL, estimated creatinine clearance (CrCL) calculated using the Cockcroft-Gault equation
<sup>a</sup> Values are mean (standard deviation) or No. (%)

### Population Pharmacokinetic Analysis

The dataset had 112 concentrations from 32 patients. Plots of dose-normalized plasma concentrations of polymyxin B versus time since last dose are presented in Figure 1. A 1-compartment model with linear elimination showed the best fit of the observed concentration-time data based on reduction in OFV and visual inspection of diagnostic plots (Figure S1). BSV was only estimated on CL, but not volume (V), in the base model, because the estimation accuracy on V was low. The proportional error model was selected to evaluate the residual variability. Covariate model building identified CrCL as the only covariate of polymyxin B CL. The plot of CrCL vs. CL of polymyxin B according to the base model is shown in Figure 2. Furthermore, there was no significant association between polymyxin B PK and age, gender, TBW, serum albumin concentration. The final PK model is shown below (Eqs. (1) and (2)).

**Figure 1.**
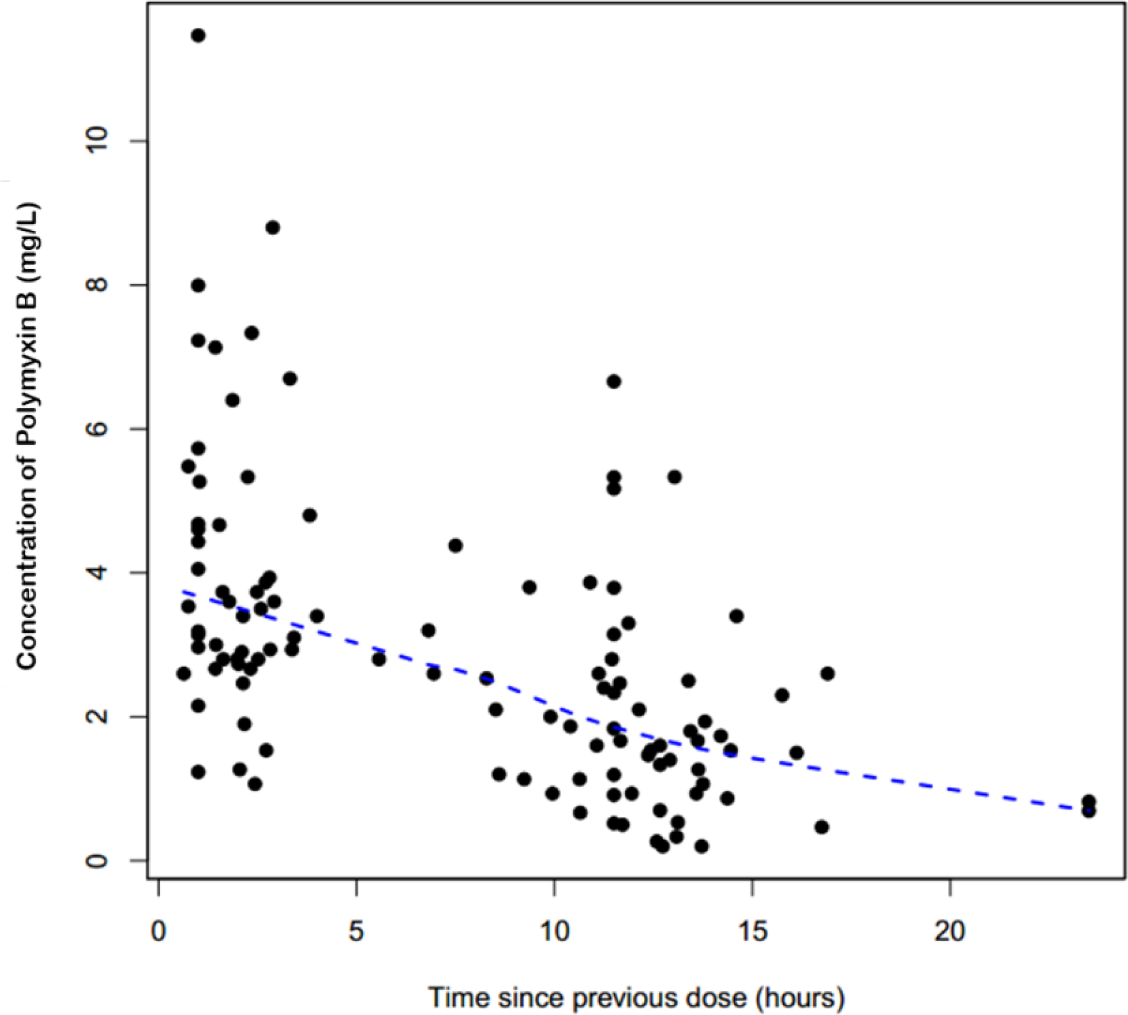
Dose-normalized serum concentration-time profiles of polymyxin B in patients.

**Figure 2.**
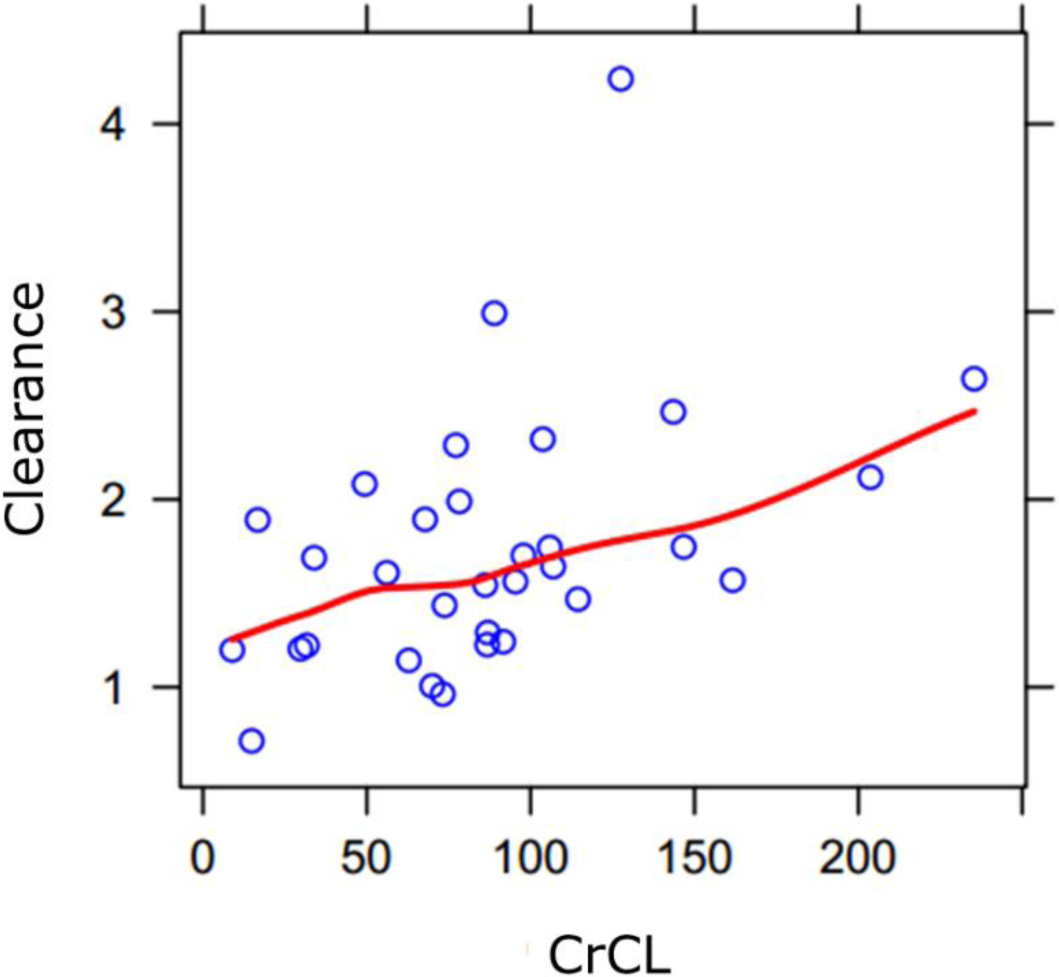
Individual polymyxin B clearance estimates versus creatinine clearance (CrCL).

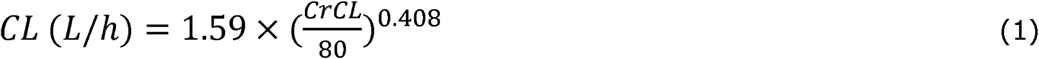

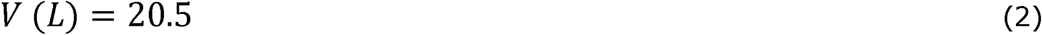

PK parameter estimates of the final covariate-structured model were shown in Table 2.

**Table 2.** Population pharmacokinetic parameter estimates from the final model.

| Parameter | Estimate | RSE% | shrinkage |
| --- | --- | --- | --- |
| **Fixed Effects** |  |  |  |
| TVCL ($\theta_1$) | 1.59 | 5.1 | |
| CrCL on CL ($\theta_2$) | 0.408 | 18.7 | |
| V ($\theta_3$) | 20.5 | 11.6 | |
| **Between-subject** |  |  |  |
| **Variability (BSV)** |  |  |  |
| BSV CL | 13% | 59.7 | 38% |
| **Residual Variability (RV)** |  |  |  |
| Proportional Error | 40.5 | 18.2 | 4% |

a. BSV calculated as $\sqrt{e^{\omega^2} - 1}$
CL covariate structure:

The goodness-of-fit plot of the final model was shown in Figure 3. The scatterplots of population prediction (PRED) and individual prediction (IPRED) revealed an improvement in the final model compared with that of basic model. The conditional weighted residuals (CWRES) vs. PRED of the final model showed a stochastic distribution around zero, and most residuals were within an acceptable range (-2 to 2). The pvcVPC of the final model showed a good fit between the observations and simulations (Figure S2). The NPDE results were shown in Figure S3 and Table S1, which showed a good performance followed a normal distribution. Mean (SD) individual empirical Bayesian estimates of polymyxin B CL was 1.75 (0.43) L/h, across all patients with V estimated at 20.5 L.

**Figure 3.**
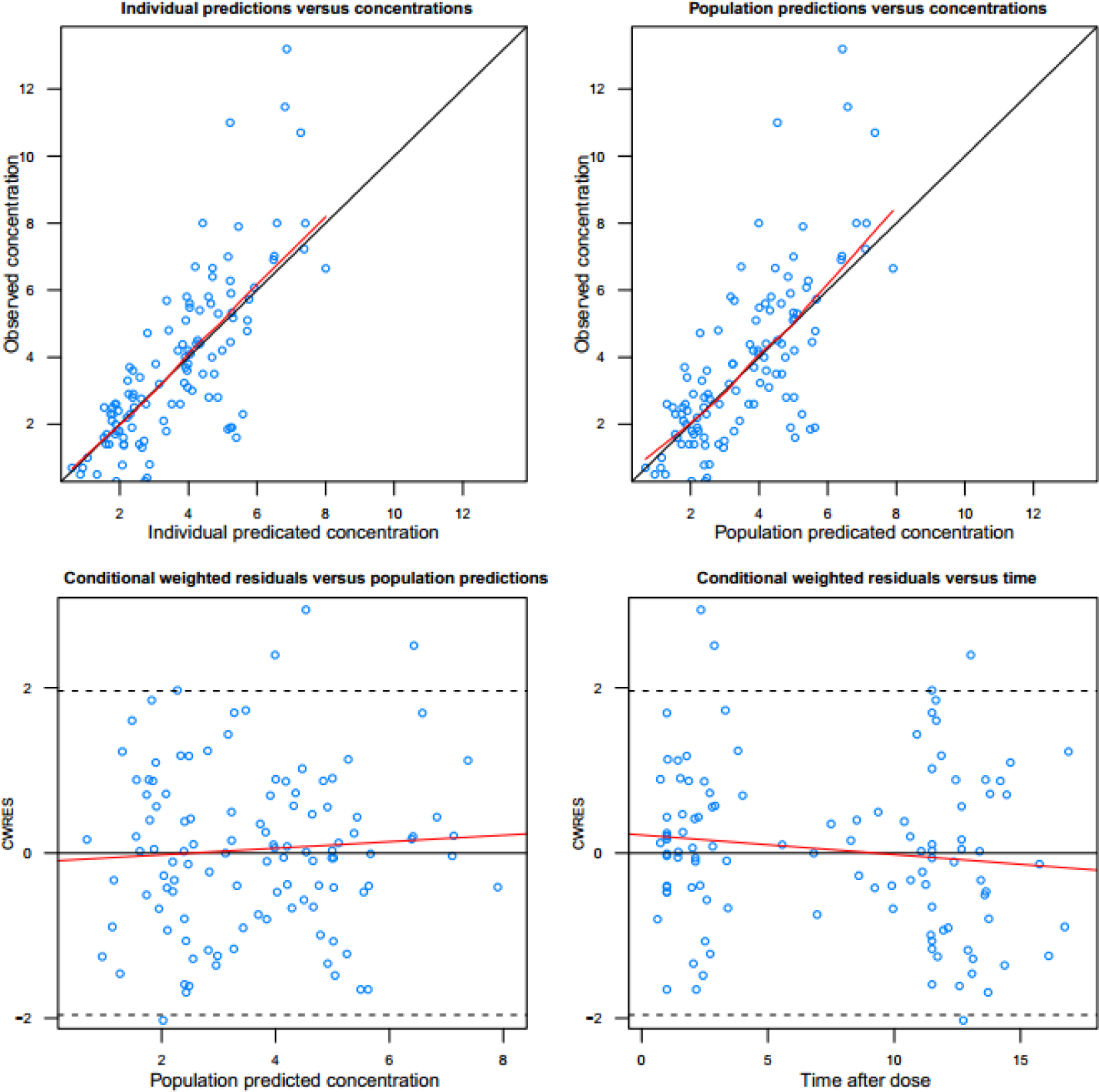
Diagnostic goodness-of-fit plots of the final model. a Observed concentration (DV) vs. individual predicted concentration (IPRED); b DV vs. population predicted concentration (PRED); c conditional weighted residuals (CWRES) vs. PRED; and d CWRES vs. time. The red lines in the upper panel represent loess smooth lines and linear fit lines, respectively.

### Monte Carlo Simulation

The AUC_ss,24h_ (AUC from 0 to 24 h) target of 50-100 mg·h/L is recommend as the PK/PD therapeutic target for maximization of efficacy for polymyxin B by the current consensus guidelines for the optimal use of polymyxins [12,18]. The lower bound of the therapeutic window was determined to be 50 mg·h/L using data from previously described murine thigh infection model studies [10,19-20]. The upper limit of the therapeutic window was determined to 100 mg·h/L based on a linear function describing the relationship between nephrotoxicity rates and polymyxin B exposures [18], in which an incidence of mild nephrotoxicity (≥ 25% decrease in creatinine clearance) of less than 40% was considered as the reasonable target. The simulated probability of achieving a target attainment of polymyxin B AUC_ss,24h_ (50-100 mg·h/L) with various dosage regimens in patients with various CrCL, predicted from Monte Carlo simulations, is quantified in Table S2 and showed in Figure 4. With the expectation of probability of target attainment (PTA) more than 90%, the recommended dosage regimen for patients with various CrCL was presented in table 3. The data showed that for patients with moderate renal insufficiency (30≤CrCL<60 mL/min), the dose should decrease 33% compared with patients with normal renal function.

**Table 3.** Recommendation of dosage regimen for patients with various patients (for MIC=1mg/L)

| **CrCL (mL/min)** | **Dosage Regimen** |
| --- | --- |
| <=20 | 30mg q12h |
| 20-30 | 40mg q12h |
| 30-70 | 50mg q12h |
| 70-120 | 75mg q12h |

**Figure 4.**
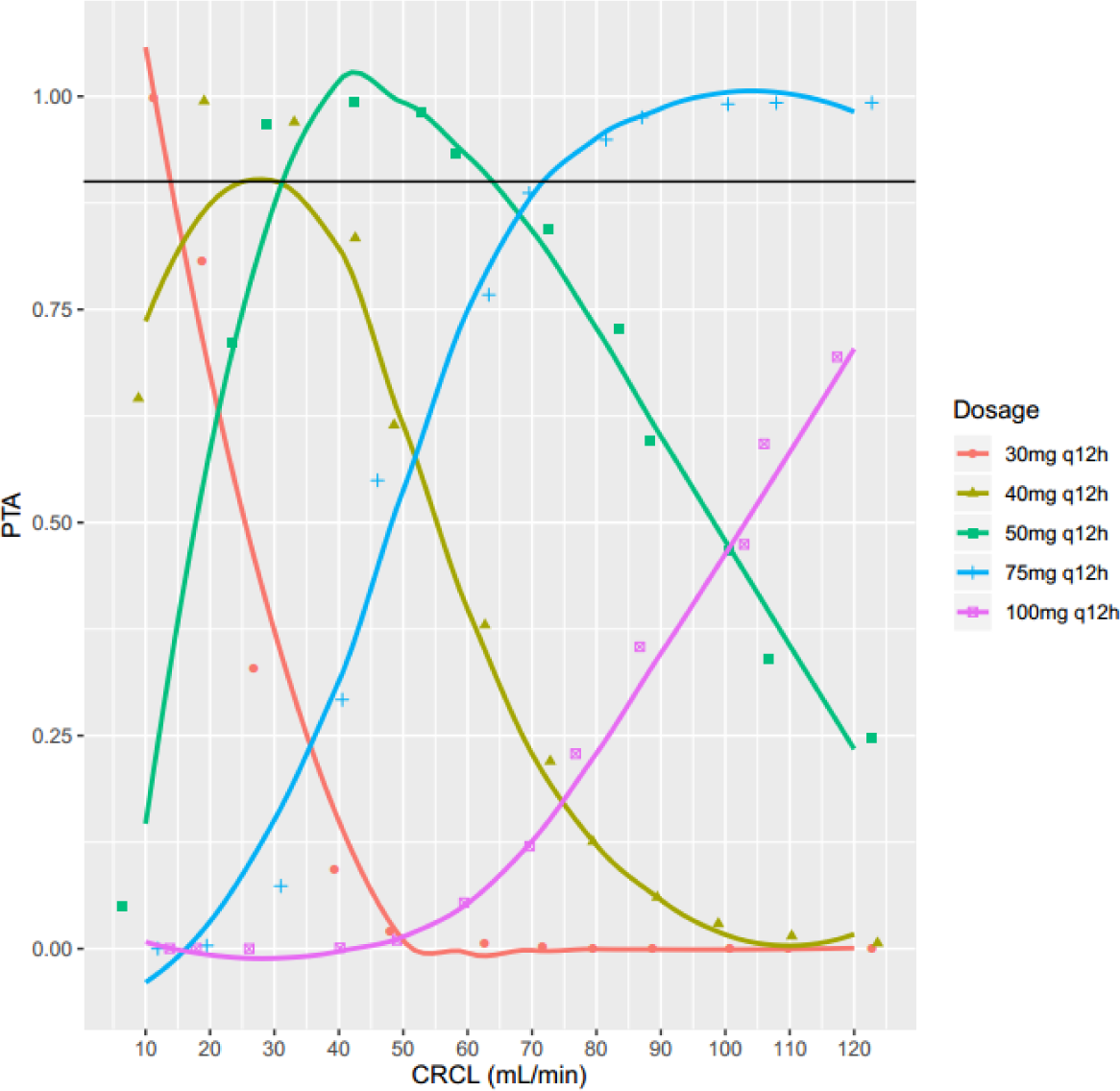
The simulated probability of achieving a target attainment of polymyxin B AUC_ss,24h_ (50-100 mg·h/L) with various dosage regimens in patients with various CrCL.

Without regard to the nephrotoxicity, the area under the concentration-time curve of free polymyxin B to MIC ratio (fAUC/MIC) is the PK/PD index that best correlates with bacterial killing [12,20]. In the thigh infection model, the *f*AUC/MIC values for 2-log bacterial killing were approximately 20 for colistin against *P. aeruginosa* and *A. baumannii* [21,22]. Considering colistin and polymyxin B have similar molecular structures and in vitro activity, it is reasonable to conclude the PK/PD of polymyxin B indices and target approach those of colistin. The protein binding of polymyxins in human plasma is ∼50% [23, 24]. Sandri et al., calculated the unbound fraction of polymyxin B in plasma in 24 patients, which showed the median unbound fraction in plasma was 0.42 [10]. Therefore, we performed Monte Carlo simulations using an unbound fraction in plasma of 0.42 to evaluate the probability of different dosage regimens reaching an fAUC/MIC of approximately 20. Only the amount of daily dose reached to 200 mg would reach to the target when the causative pathogen MIC is 2 mg/L (Table S3 and Figure 5). However, 200 mg daily dose would increase the risk of nephrotoxicity (Figure 4). Thus, for severe infections caused by organisms with polymyxin B MIC of ≥ 2 mg/L, though a high daily dose (e.g. 200 mg/day) would possible for bacterial eradication, the risk of nephrotoxicity is significantly increased.

**Figure 5.**
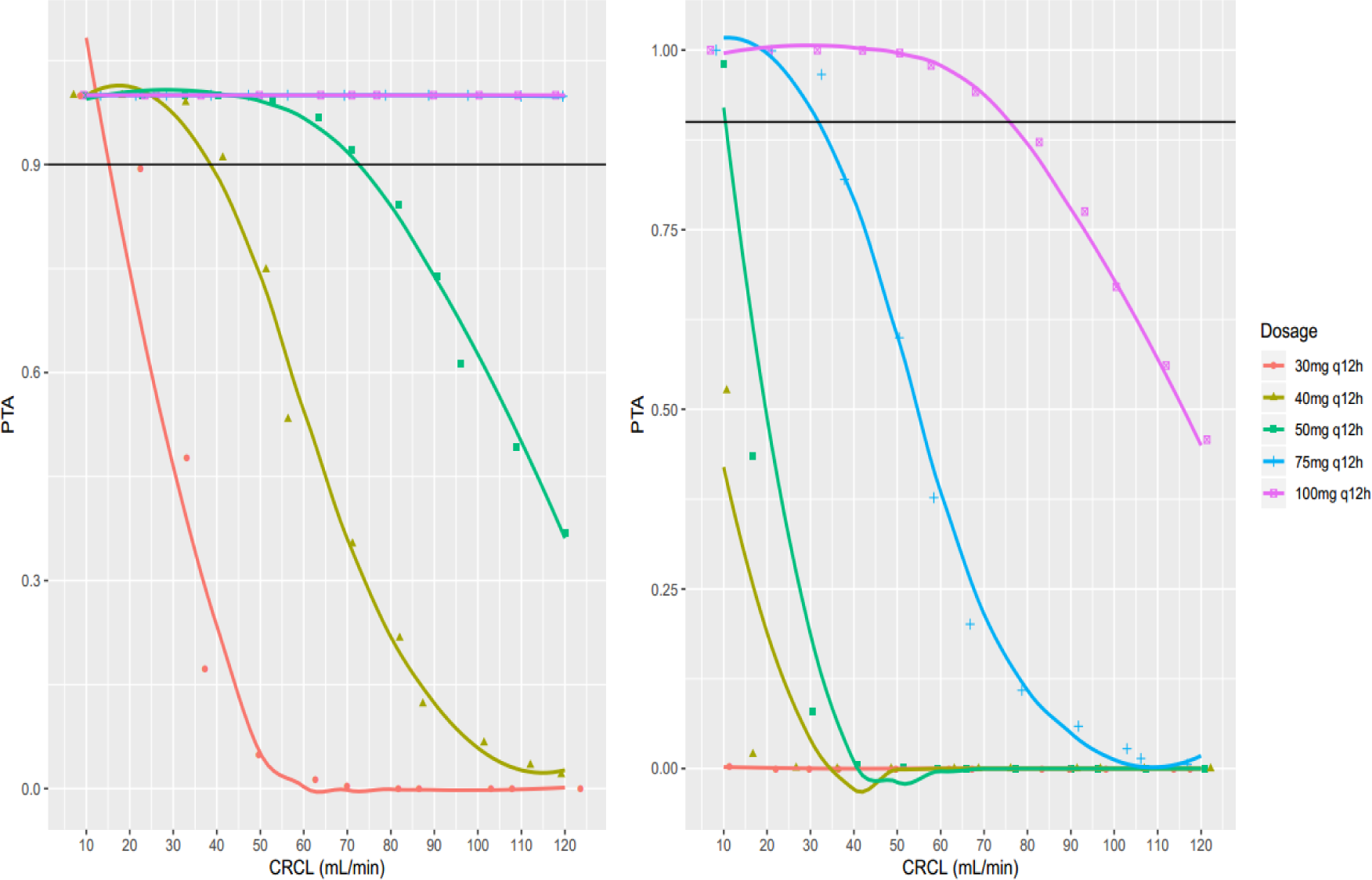
The simulated probability of achieving a target attainment of polymyxin B fAUC_ss,24h_/MIC≥ 20 (MIC=1 mg/L (left), MIC=2 mg/L (right)) with various dosage regimens in patients with various CrCL.

## Discussion

This study has made a significant contribution to understanding of how to optimize the clinical use of polymyxin B in patients with various CrCL. It is the first study to data using the real-world data from routine therapeutic drug monitoring built a population pharmacokinetics model, which showed a significant association between CrCL and polymyxin B CL, suggest dose reducing in patient with renal insufficiency, which is in agreement with the recommendation provided by current FDA approved package of polymyxin B. We also provide the dosage regimens for patients with various renal function according to the recommended PK/PD therapeutic target for maximization of efficacy for polymyxin B.

It was commonly recommended in the package inserts for polymyxin B that the dose should be reduced in patients with renal insufficiency. However, data from several recent studies have contradicting insights refuting this recommendation [10-11, 25-26] that polymyxin B is not significantly eliminated by the kidneys and polymyxin B clearance does not depend on CrCL. Therefore, the PK rational for not adjusting doses according to renal function was concluded. More PK studies in patients with renal insufficiency are needed to validate the PK rational for not adjusting the dose in patient with renal insufficiency, which contradicts the recommendation from the package.

According to our final model, the inclusion of CrCL significantly improved the goodness-of-fit of the model. The data used for Pop PK analysis included 15 patients with normal renal function (estimated creatinine clearance (CrCL)≥80mL/min) at baseline and 17 patients with renal insufficiency (CrCL< 80mL/min). The creatinine clearance in patients was 5.91-244 mL/min, in which 3 patients was with extremely lower serum creatinine below 35 μmol/L. Therefore, the percentage of patients with or without renal insufficiency is similar, while the distribution of renal function was extensive in this study. In accordance with our results, Avedissian and colleagues [27] found a potential relationship between the CrCL and polymyxin B CL in 9 cystic fibrosis patients, which was best described via a Hill function. In addition, polymyxin B’s proposed mechanisms of elimination involve both renal (via renal tubular reabsorption) and non-renal pathways. The potential reason for the lower total clearance of polymyxin B in patients with renal insufficiency might be that the glomerular filtration function in these patients is impaired, which caused reduced amount of polymyxin B being filtrated.

Despite the renewed interest in polymyxin B used for treatment of multidrug resistant Gram-negative bacteria, the optimal dosage strategy is still unclear because they are primarily based on population pharmacokinetics. The packages insert for polymyxin B recommended initial dosing polymyxin B via intravenous way at 1.5-2.5 mg/kg/d, divided into two doses [13]. The data of Monte Carlo simulation showed that for patients with moderate renal insufficiency (30≤CrCL<60 mL/min), the dose should decrease 33% compared with patients with normal renal function. Therefore, physicians should balance the risk of polymyxin B-induced nephrotoxicity against the benefit of adequate doses of polymyxin B in patients with declining renal function. In addition, when the causative pathogen MIC is 2 mg/L, though dosage regimen of 100 mg q12h could reach an fAUC/MIC more than 20 at steady state, the risk of nephrotoxicity was incredibly increased. For pathogens with MICs of 2 mg/L, only a very small proportion of patients will reach the therapeutic target of AUC_ss,24h_ (50-100 mg·h/L).

A potential limitation of our study was the data that used for PK analysis was retrospectively collected, despite the time of dosing and sampling were precisely recorded. In addition, though the relation between renal function and polymyxin B CL was quantificationally studied, the confirmation of urinary excretion of polymyxin B should be further investigated.

In conclusion, this is the first study to demonstrate that the CrCL is a patient characteristic that influences polymyxin B PK and provide the dosage regimens for patients with various renal function according to the therapeutic target of AUCss,24h (50-100 mg·h/L). Dose reducing downward of polymyxin B according to CrCL is recommended for individuals with renal insufficiency. In addition, for severe infections caused by organisms with polymyxin B MIC ≥ 2 mg/L, though a high daily dose (e.g. 200 mg/day) would possible for bacterial eradication, the risk of nephrotoxicity is significantly increased.

## Data Availability

The Data are available for inquiry

## Acknowledgements

The contents of the manuscript are solely the responsibility of the authors and all authors have no actual or potential conflicts of interest to report.

